# Human mobility and COVID-19 transmission: a systematic review and future directions

**DOI:** 10.1101/2021.02.02.21250889

**Authors:** Mengxi Zhang, Siqin Wang, Tao Hu, Xiaokang Fu, Xiaoyue Wang, Yaxin Hu, Briana Halloran, Yunhe Cui, Haokun Liu, Zhimin Liu, Shuming Bao

## Abstract

Without a widely distributed vaccine, controlling human mobility has been identified and promoted as the primary strategy to mitigate the transmission of COVID-19. Many studies have reported the relationship between human mobility and COVID-19 transmission by utilizing the spatial-temporal information of mobility data from various sources. To better understand the role of human mobility in the pandemic, we conducted a systematic review of articles that measure the relationship between human mobility and COVID-19 in terms of their data sources, statistical models, and key findings. Following the guidelines of the Preferred Reporting Items for Systematic Reviews and Meta-Analyses (PRISMA) statement, we selected 47 articles from Web of Science Core Collection up to September 2020. Restricting human mobility reduced the transmission of COVID-19 spatially, although the effectiveness and stringency of policy implementation vary temporally and spatially across different stages of the pandemic. We call for prompt and sustainable measures to control the pandemic. We also recommend researchers 1) to enhance multi-disciplinary collaboration; 2) to adjust the implementation and stringency of mobility-control policies in corresponding to the rapid change of the pandemic; 3) to improve statistical models used in analyzing, simulating, and predicting the transmission of the disease; and 4) to enrich the source of mobility data to ensure data accuracy and suability.

## 1. Introduction

Human mobility plays an important role in the transmission of infectious diseases. With the increase of human mobility caused by the development of transportation networks and globalization, the spread of infectious diseases can be unprecedentedly rapid and difficult to prevent and control, resulting in pandemics. Such pandemics have been witnessed in history, for example, the 1918 novel influenza A (H1N1) pandemic, the 2009 H1N1 pandemic, and the current coronavirus disease 2019 (COVID-19). Without a widely distributed vaccine, controlling human mobility has been identified and promoted as the primary strategy to mitigate the transmission of COVID-19 (Gatto, Bertuzzo et al. 2020, Kraemer, Yang et al. 2020, Wang, Liu et al. 2020, Yabe, Tsubouchi et al. 2020). During this pandemic, various policies have been implemented worldwide to restrict human mobility across and within countries including international travel bans and national border closures, restrictions between states and cities, stay-at-home orders, limited private and public gatherings, in addition to closing schools, universities, workplaces, and public transportation (Hale and Webster 2020).

Since the outbreak, academic researchers have put substantial efforts into studying the relationship between human mobility and COVID-19 transmission (hereinafter referred to as “mobility-transmission relationship”). Many studies have reported the efficacy of mobility restrictions on controlling the spread of the virus (Kraemer, Yang et al. 2020, Wang, Liu et al. 2020, Yabe, Tsubouchi et al. 2020). However, the timeline and stringency of social restriction policies and lockdown orders have been vociferously challenged due to significant social and economic costs (Bonaccorsi, Pierri et al. 2020, Lecocq, Hicks et al. 2020). In addition, the scientific comparison of data and methodologies used to examine the mobility-transmission relationship has been under-explored.

To fulfill the aforementioned knowledge gaps, we conducted a systematic review of articles that measure the mobility-transmission relationship in terms of their study purposes, data usage, statistical models, and key findings. Based on our review, we offered future research directions in the spatial and temporal dimensions to researchers with similar interests in this topic. Through collective efforts from multiple disciplines, we hope to mitigate the spread of infectious diseases with evidence-based solutions and to be better prepared for the potential outbreak of future infectious diseases given the increased globalization, urbanization, and interruption of human beings to eco-systems.

## 2. Method

We followed the guidelines of the Preferred Reporting Items for Systematic Reviews and Meta-Analyses (PRISMA) statement to select articles and to report the findings. PRISMA statement is a guideline developed to support researchers to conduct systematic reviews (Moher, Liberati et al. 2009). We applied the checklist of PRISMA with the items to report in a systematic review and a flow diagram indicating the workflow of selecting articles in a systemic review (Moher, Liberati et al. 2009). We commenced with searching through the Web of Science (WoS) Core Collection of all the published articles between January 2020 to September 2020 to cover the most recent publications with the topic of human mobility and COVID-19. WoS is the most widely used and authoritative database of research publications and citations. WoS Core Collection Coverage includes more than 20,900 journals plus books and conference proceedings from various disciplines (Birkle, Pendlebury et al. 2020). The searching terms we used are “((COVID-19 OR “novel coronaviruses” OR 2019-nCov OR SARS CoV-2) AND (“human mobility” OR “human movement” OR “population flow” OR “social distanc*” OR “physical distanc*” OR “travel restriction” OR “movement control” OR stay-at-home OR lockdown OR shelter-in-place))”.

The flow diagram of the article selection through different phases of the review was presented in Figure 1. We limited our search to published and early access articles, resulting in a total number of 1,649 articles. We then excluded the articles in irrelevant areas (e.g., psychology, neuroscience, neurology, and surgery), narrowing down 868 articles. We further excluded 755 articles that do not meet our inclusion criteria (Table 1) by screening their titles and abstracts. Through reading and assessing full texts, 47 articles highly relevant to our review’s interest were finally selected. We summarized and analyzed the information of the study countries/regions, study purposes, data resources, statistical models, and key findings. This information was summarized in Supplementary Table 1. Based on what we found, we proposed future research directions of mobility-transmission studies.

**Table 1.** Inclusion and exclusion criteria of article selection

| **Criteria** | **Definition** |
| --- | --- |
| Inclusion | 1) measure the association between human mobility and COVID-19 transmission, 2) measure the association between human mobility-related policies and COVID-19 transmission, and 3) apply quantitative methods. |
| Exclusion | 1) measure the association between human mobility and other COVID-19 pandemic related issues, e.g., economic development, mental health, family relationship, 2) apply qualitative methods. |

**Figure 1.**
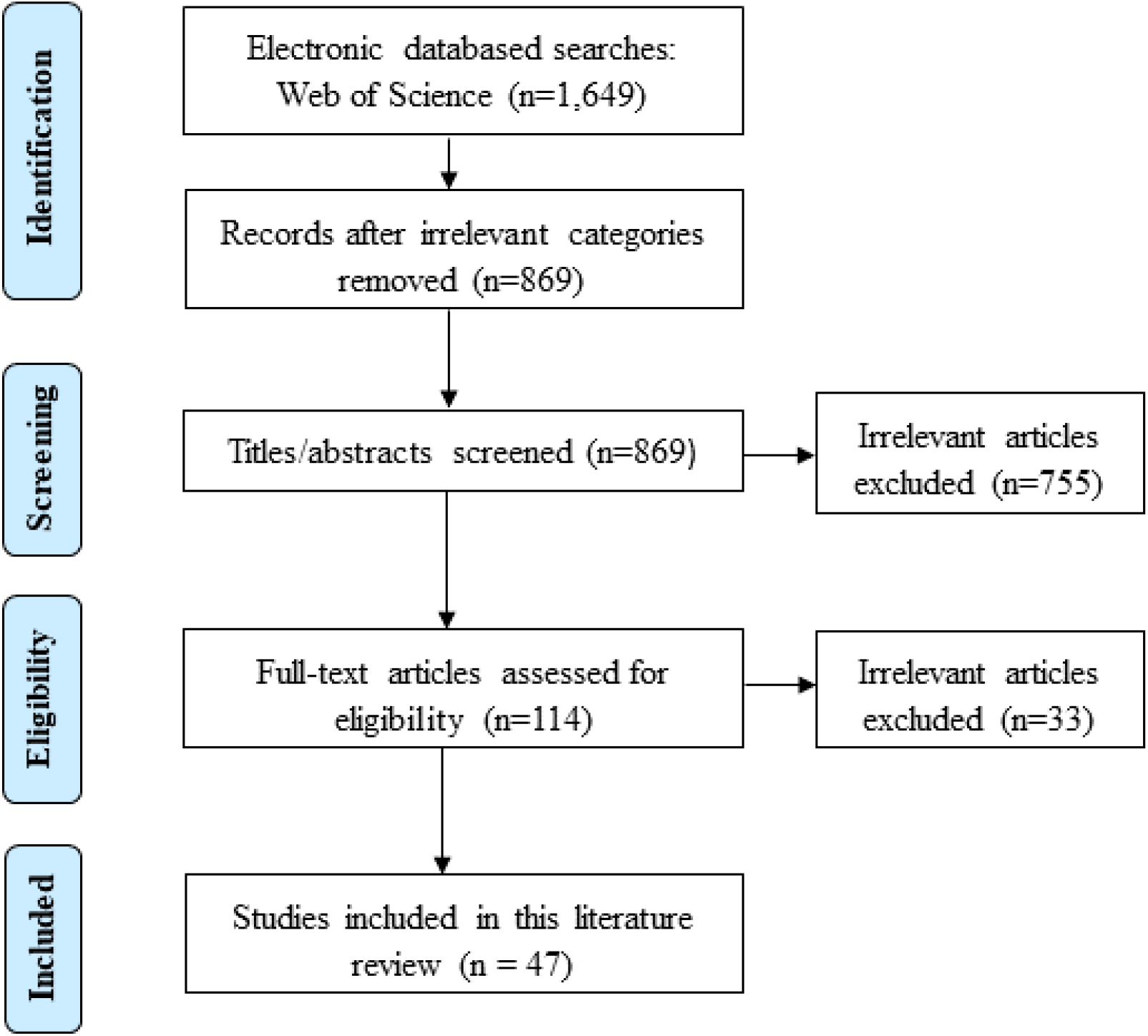
PRISMA flow chart on the identification and screening of studies on human mobility and COVID-19 transmission

**Supplementary Table S1.**
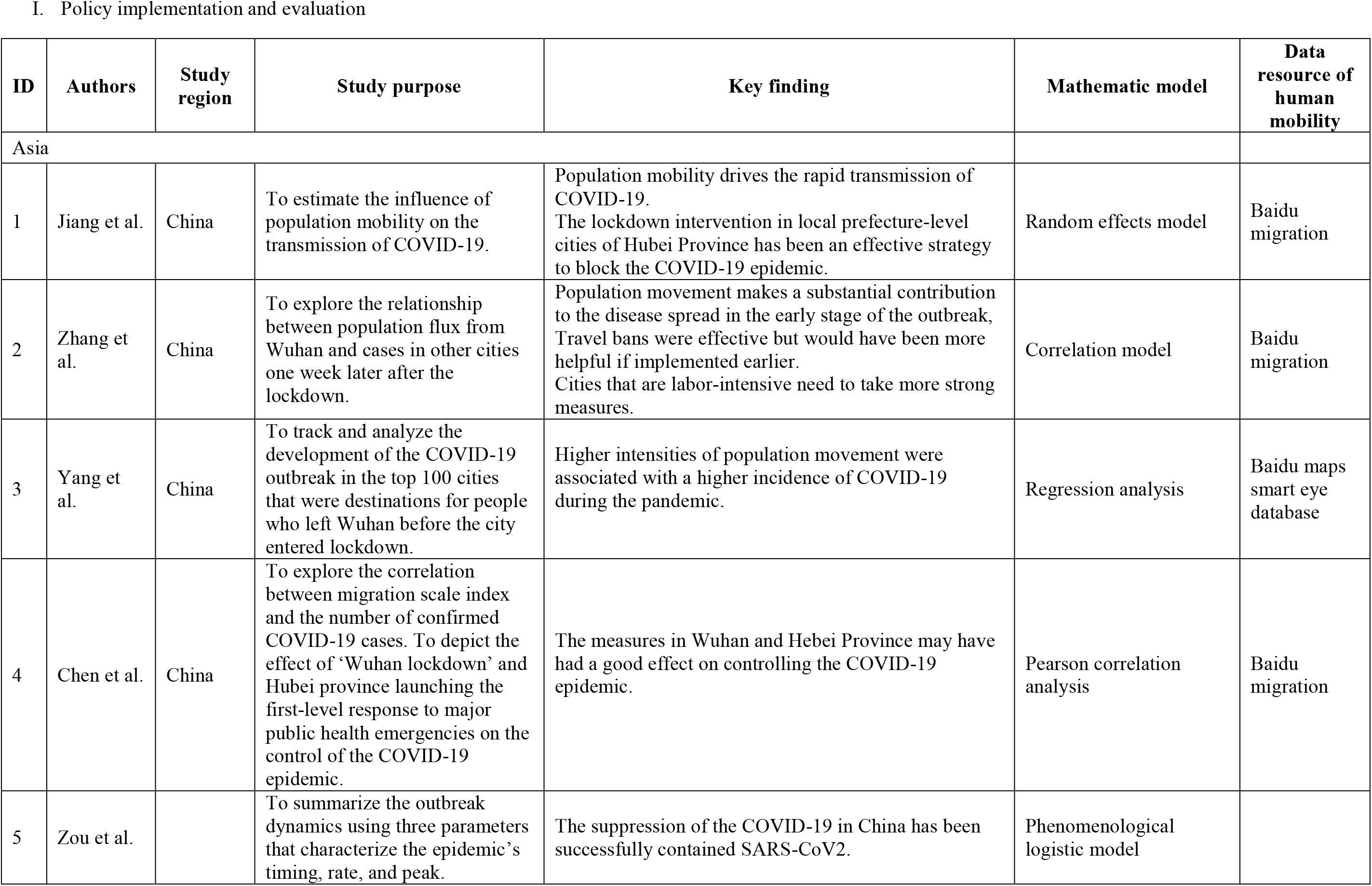
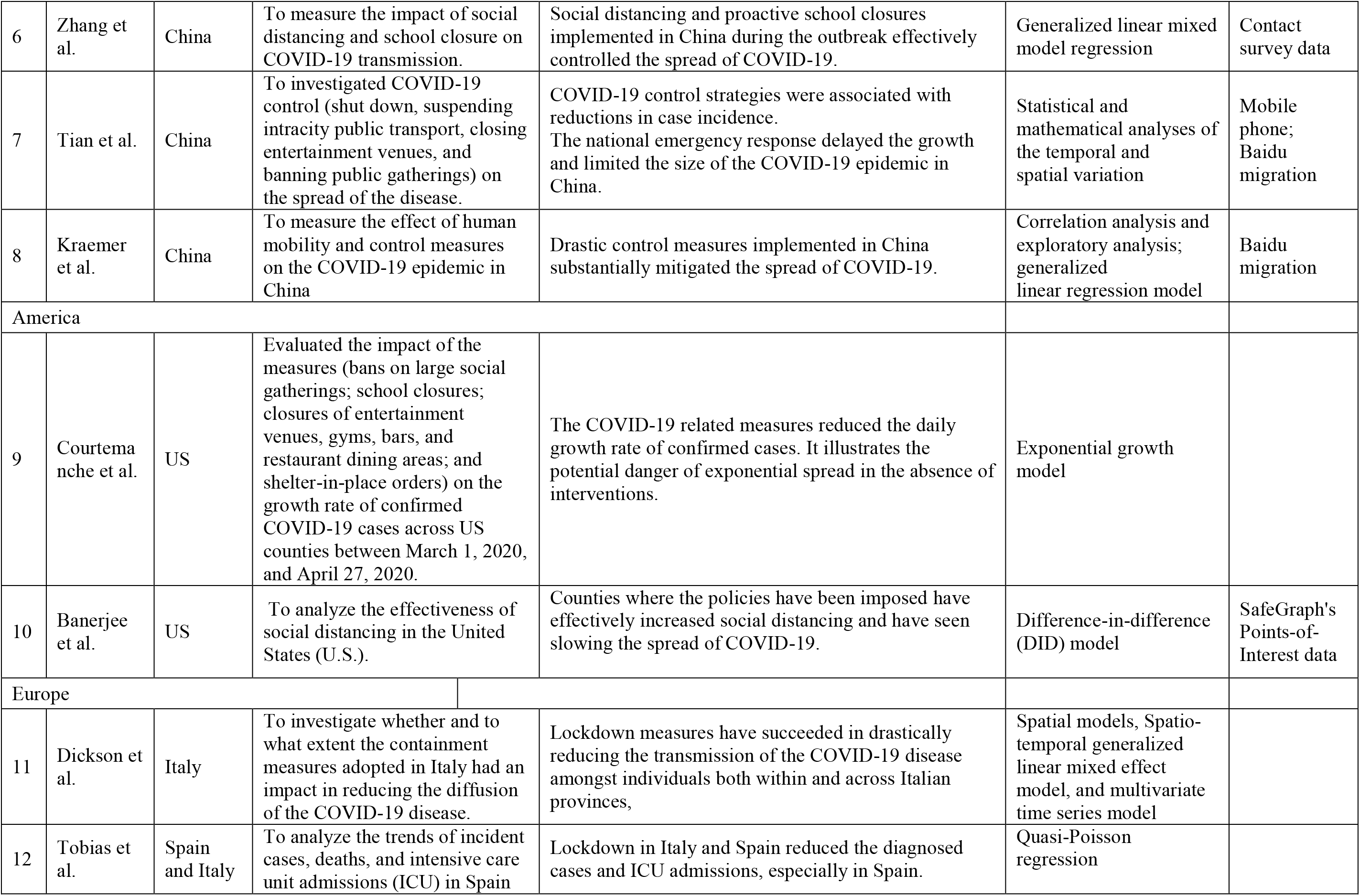
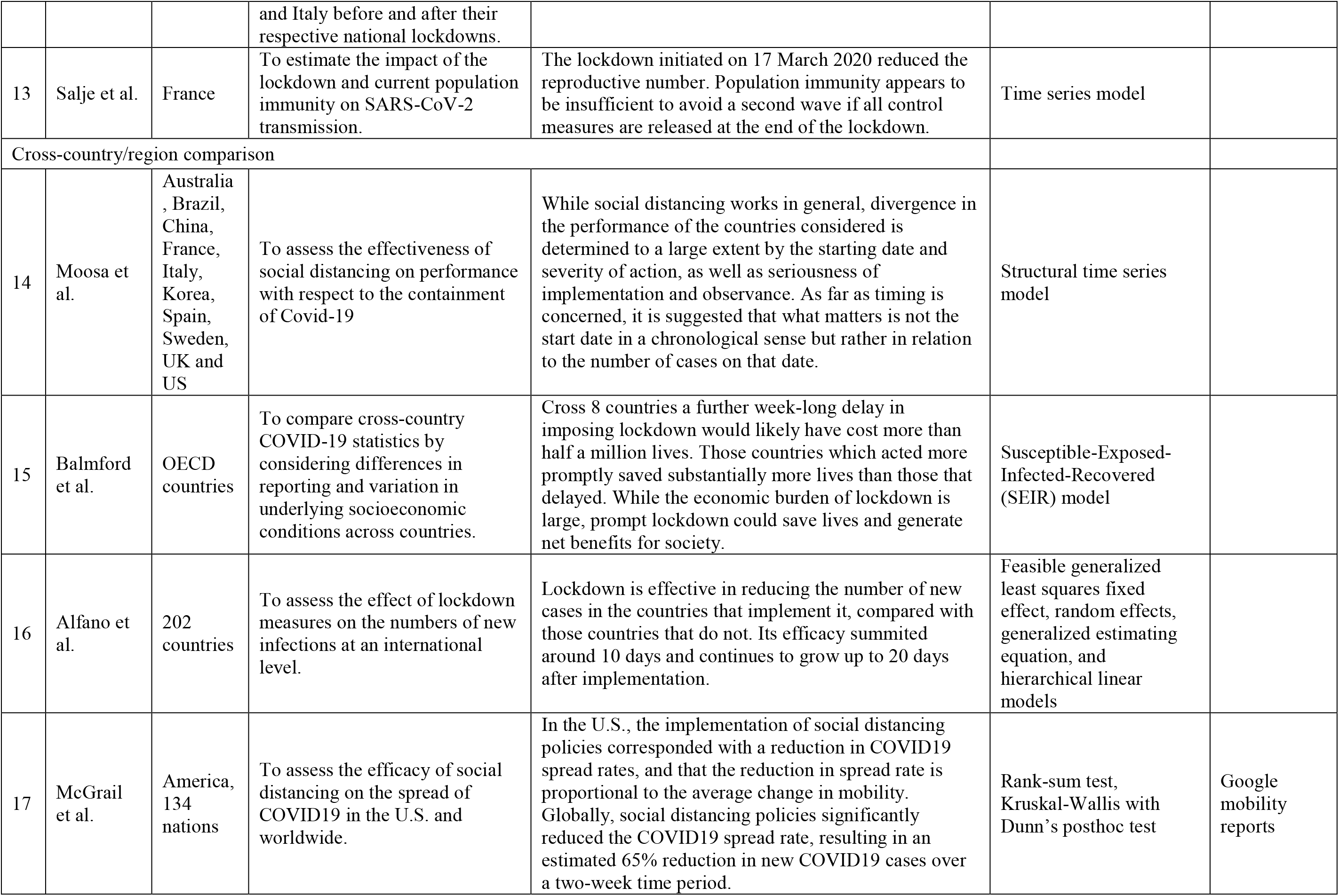
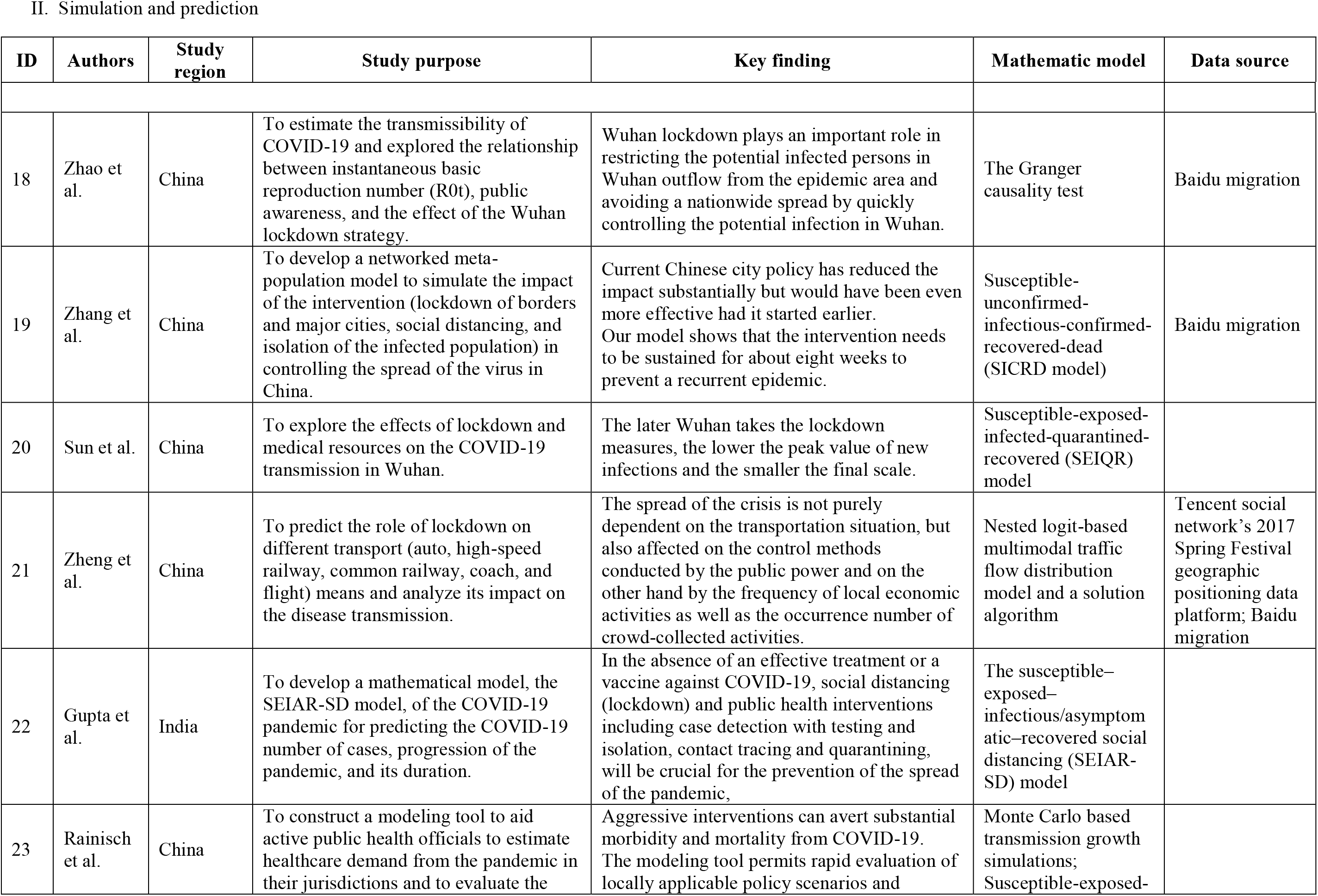
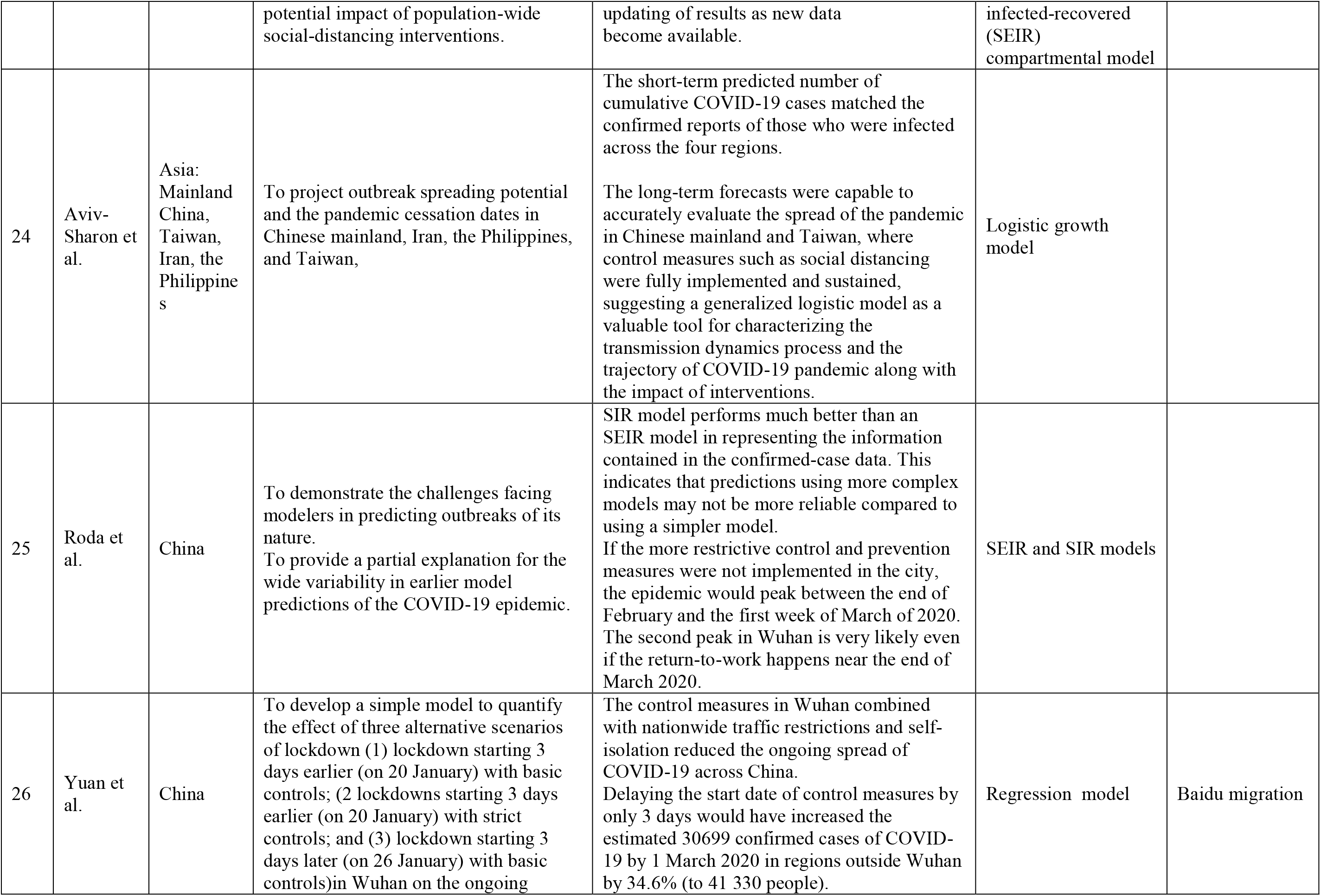
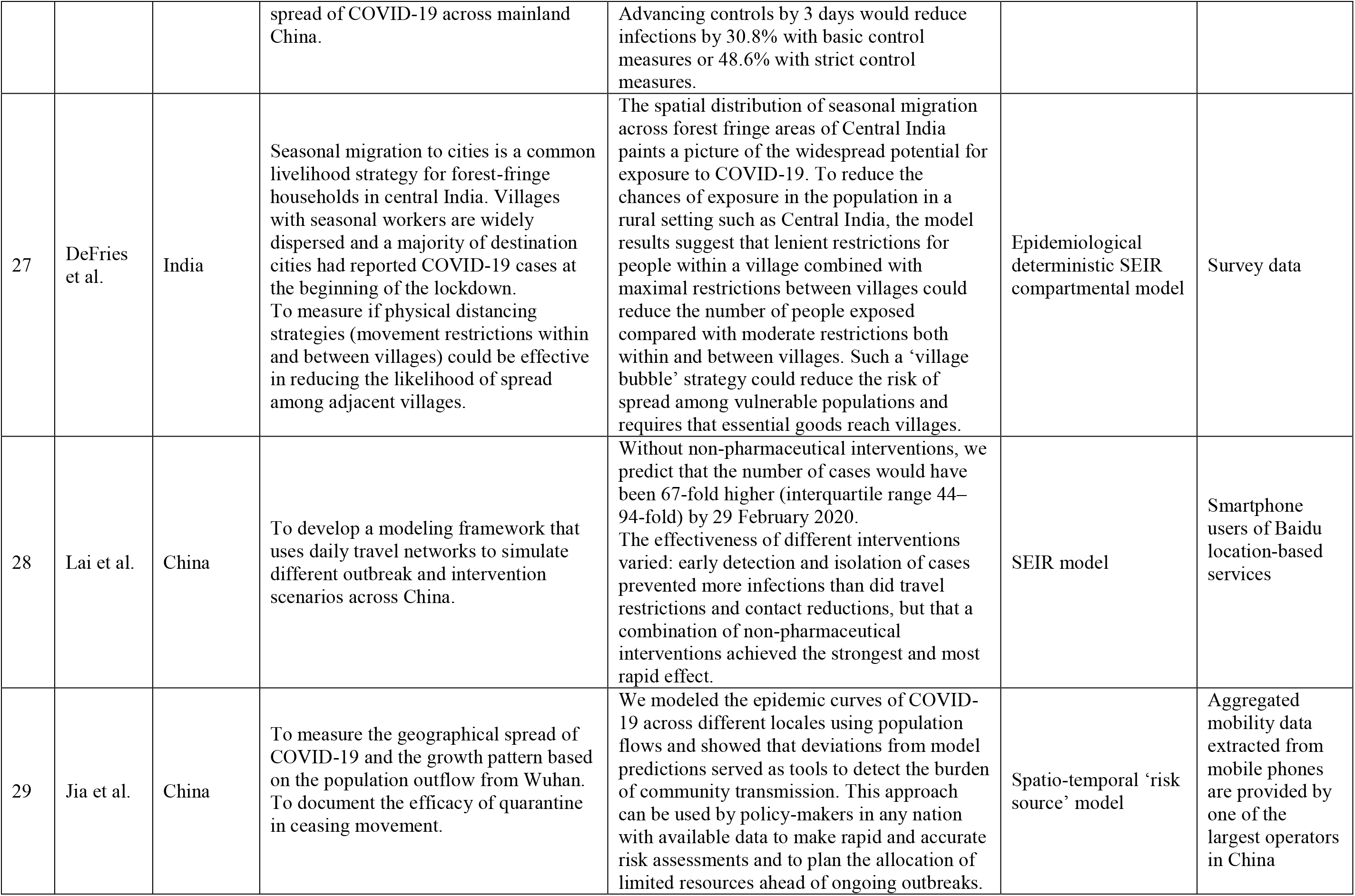
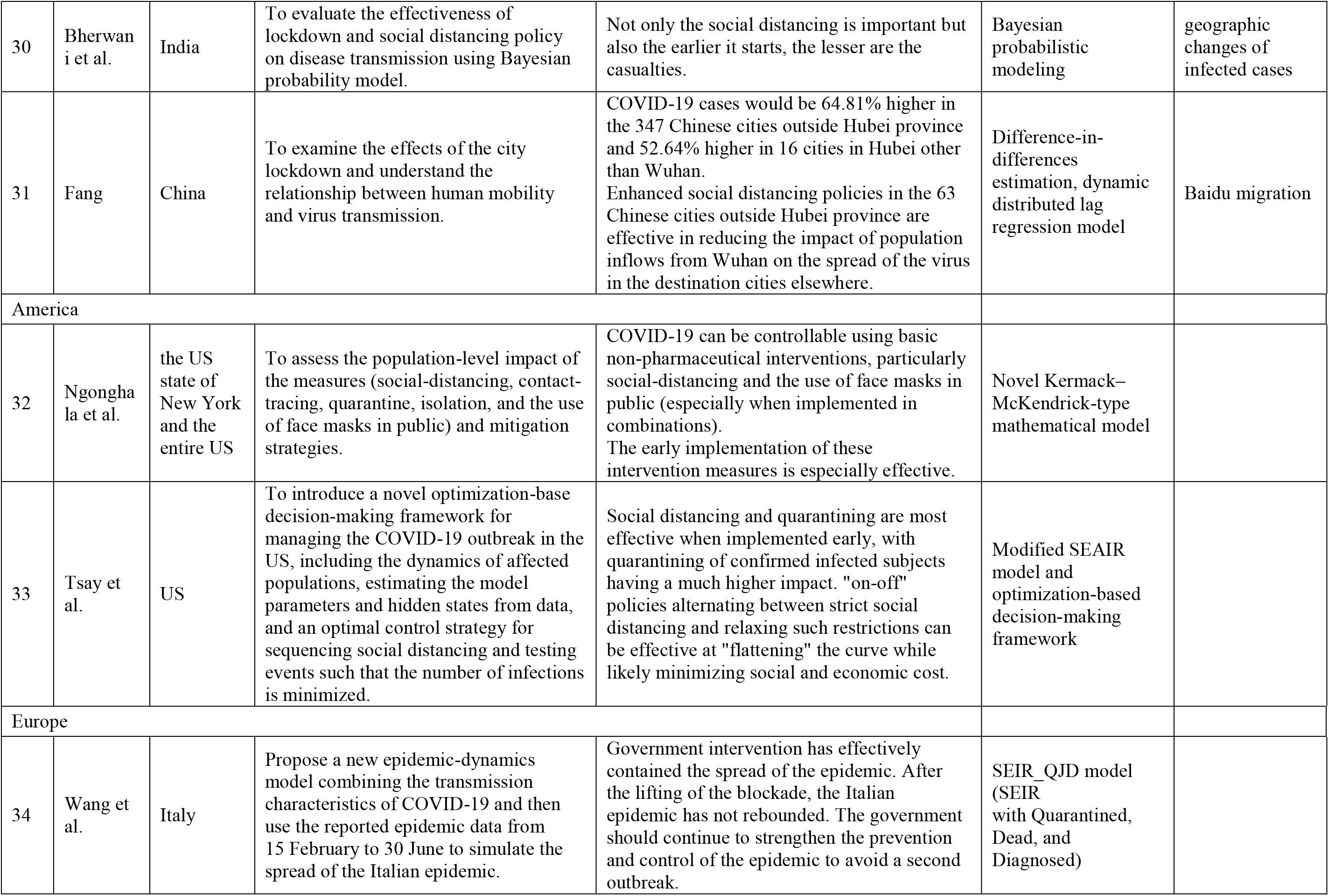
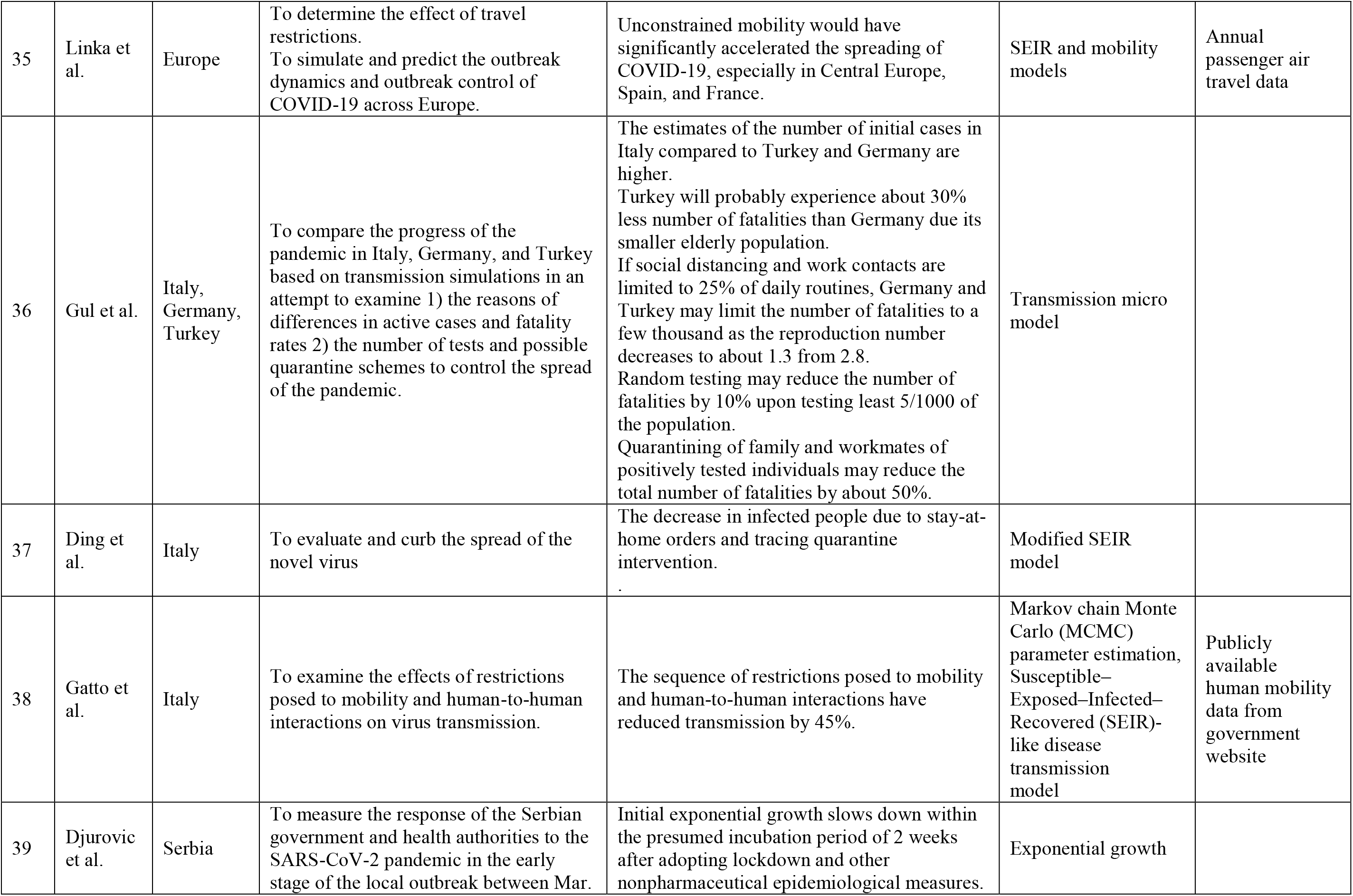
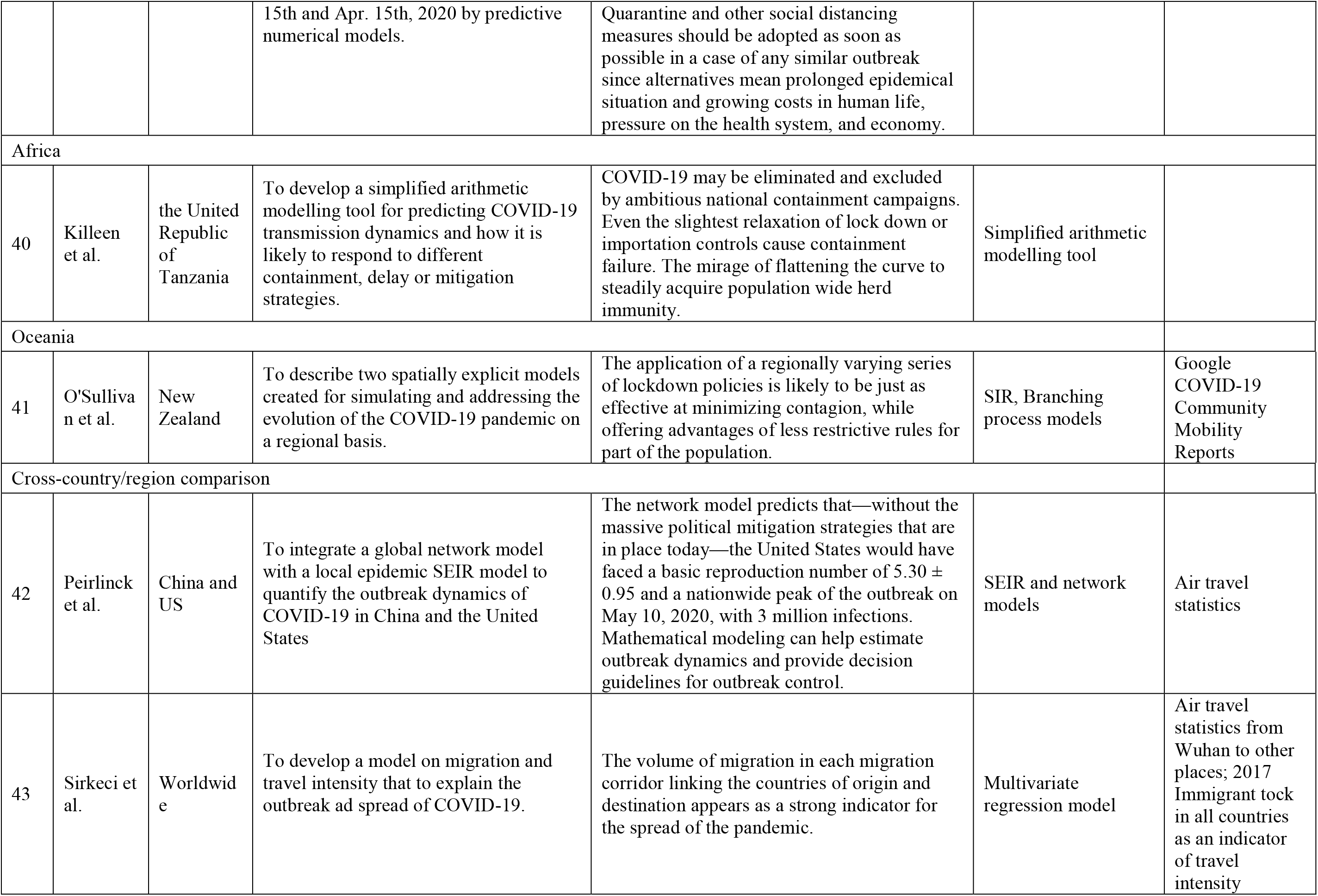
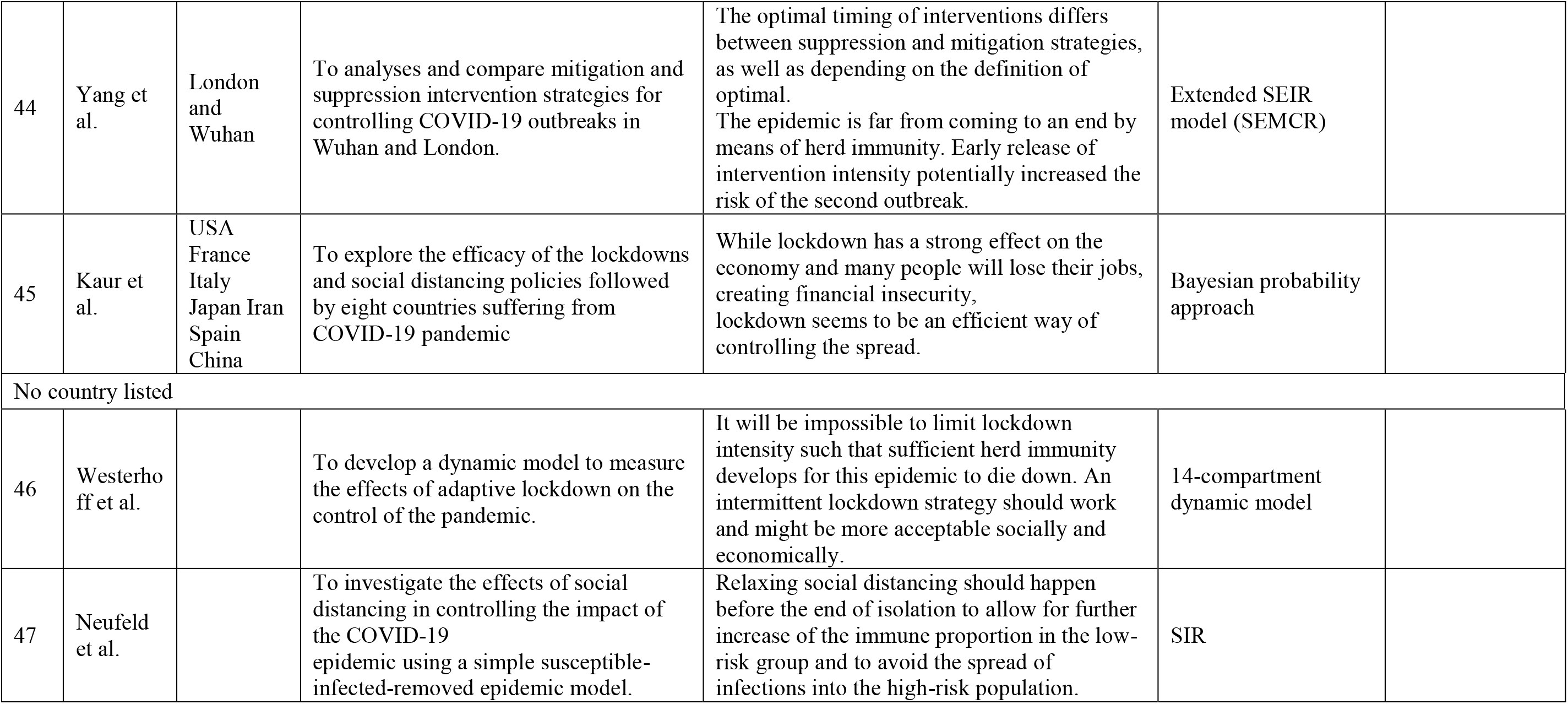
Characteristics of the selected articles based on its study purpose and study region

## 3. Results

### 3.1. Data sources and features

The selected papers mainly rely on two types of data: COVID-19 data at different scales and human mobility data. COVID-19 data in terms of the number of confirmed and susceptible cases, deaths, and recovered cases are usually easy to retrieve from research institutes, public health authorities, or government reports, while human mobility data are multi-sourced with specific applications. This review mainly focuses on human mobility data in terms of its sources, public accessibility, spatial coverage, time coverage, update frequency, advantages, and disadvantages (Table 2).

**Table 2.**
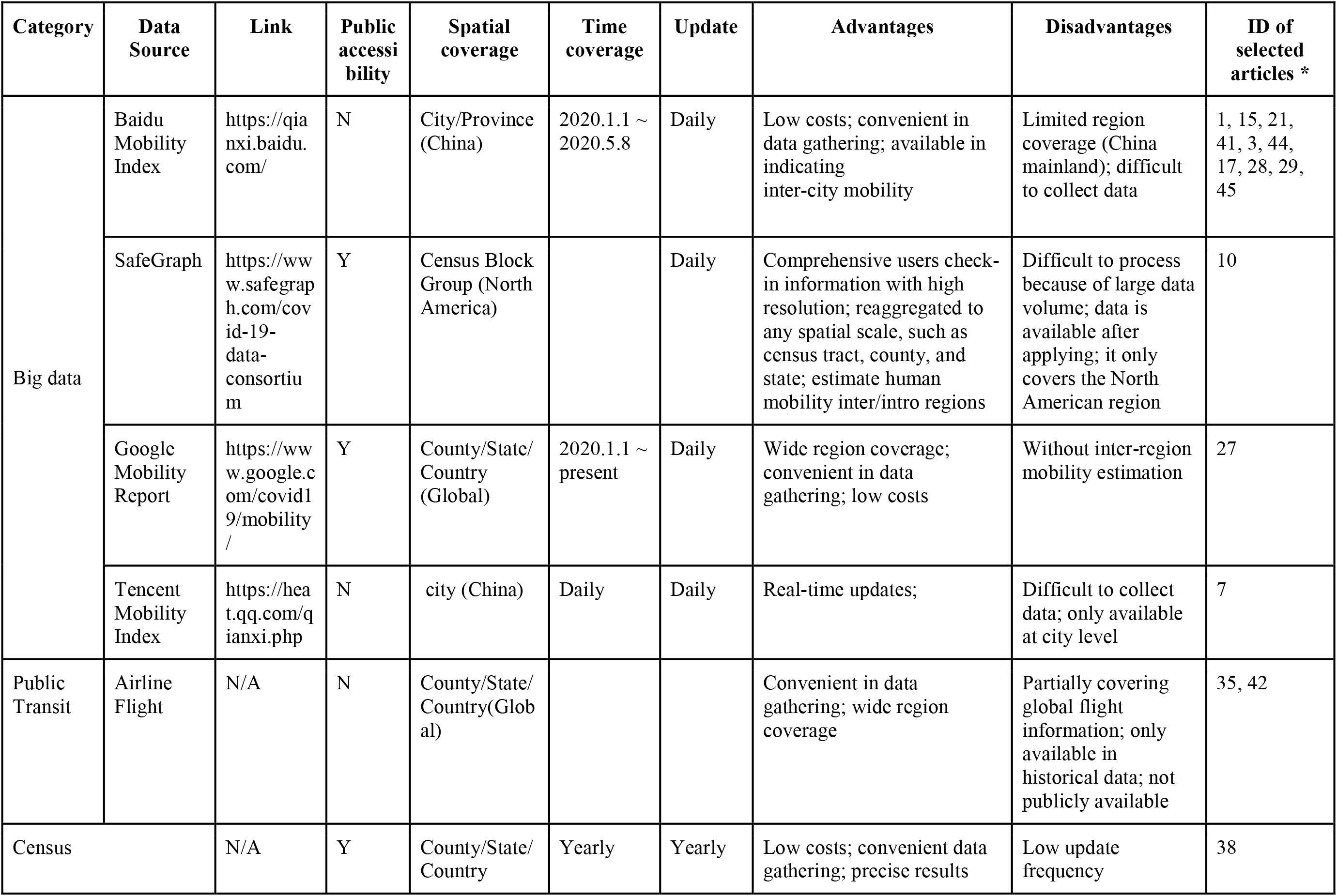
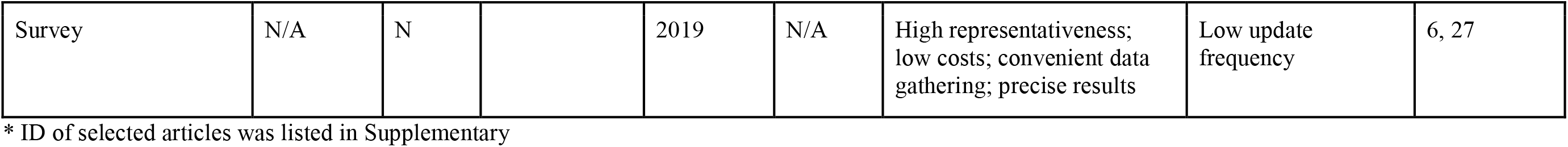
Summary of mobility data sources used in the selected articles

The first type of mobility data is big data gathered by technology companies, including Baidu, SafeGraph, Google, and Tencent. For example, Google mobility data is created by aggregated and anonymized data sets from users who have turned on the *Location History* setting in the products such as Google Maps (Aktay, Bavadekar et al. 2020). It is encoded as a percentage change in the mobility metric compared to the baseline of human mobility from January 3 to February 6, 2020 (Aktay, Bavadekar et al. 2020). Unlike Google mobility data, SafeGraph mobility data adds unique and valuable insights into the mobility change by estimating the aggregated and anonymized summary of foot traffic to 6 million points-of-interest in North America (Safegraph 2021). Safegraph aggregates the data by category (e.g., Airports or Supermarkets) or brands (e.g., Costco or McDonald’s) (Safegraph 2021). In addition, Baidu, a Chinese leading information technology (IT) company, offers location-based services to mobile devices for online searching and mapping based on the Global Positioning System, Internet Protocol addresses, locations of signaling towers, and wireless networks. Baidu Mobility Index contains daily inbound (i.e., percentage of people traveling to the city from all the cities in China) and outbound (i.e., percentage of people traveling from the city to all the cities in China) mobility data for all cities in China (except for Hong Kong, Macau and Taiwan) on each day from January 1, 2020, to May 7, 2020 (Liu, Clemente et al. 2020). Baidu mobility data has been widely used to study how the population migration at the early stage of the COVID-19 outbreak in China. Similarly, Tencent is another Chinese leading IT company providing inter-city human mobility information by integrating air flight, train, and vehicle data. However, Tencent only released the inflow and outflow data from10 Chinese cities with the highest mobility index.

Advantages of big data in human mobility include timeliness, cost-effectiveness, and large spatial coverage, while its disadvantages vary across different data sources. For example, Google mobility data covers most countries worldwide, while Baidu and Tencent mobility data only covers Mainland China. Compared with Google mobility data, Baidu mobility data is relatively difficult to retrieve, requesting users to develop programs to access data, and the available data is restricted for a certain period. However, Google mobility data cannot indicate the inter-regional mobility flow.

The second type of mobility data is the public transit data, collected through air flights. For example, the Bureau of Transportation Statistics in the U.S. published passenger numbers of flights at the state and national level (United States Department of Transportation), which was used by Peirlinck et al. to model the spreading of COVID-19 across the U.S. (Peirlinck, Linka et al. 2020). Public transit data has the advantage of compensating for the international or inter-regional mobility estimates, which cannot be revealed by Google mobility data. However, its key disadvantage is the roughness and availability at a relatively coarse scale, which cannot accurately simulate the spreading of COVID-19 at a fine scale.

The third category is census data, which records the number of people moving between or within administrative regions. For example, the United States Census Bureau publishes yearly geographic mobility dataset by region and category including race, sex, age, relationship to householder, educational attainment, marital status, nativity, tenure, and poverty status at the national, inter-state, intra-state, inter-county, and intra-county level (United States Census Bureau). When building epidemic models to estimate the effect of human mobility on COVID-19 transmission, Gatto et al. identified mobility fluxes at the municipal and provincial levels based on the 2011 commuting data from the Italian Census Bureau (Gatto, Bertuzzo et al. 2020). Census data is representative, easy to access, and usually available at various spatial scales (e.g., county, states, and nationwide). However, census data is usually updated infrequently, its data could be too out-of-date and unable to reflect changes in human mobility with the rapid response to the implementation of social restriction and lockdown policies during the pandemic.

The fourth category is survey data, containing the participants’ personal and location information. Such survey data collects accurate and verifiable data based on the survey questions. For example, DeFries et al. identified migration patterns over the last five years using a collected household survey covering 5,000 villages in 32 districts in central India (DeFries, Agarwala et al. 2020). Zhang et al. analyzed 1,245 contacts reported by 636 survey participants in Wuhan and 1, 296 contacts reported by 557 survey participants in Shanghai to study the impact of social distancing and school closure on COVID-19 transmission. As a traditional source to track human mobility, survey data is relevant to the study objective. It could cover those who are eliminated in secondary data resources. However, it is time-consuming and expensive for data collection, especially to collect a large number of participants from various regions. Thus, it is mainly used to measure human mobility in relatively small areas compared with the other types of data.

### 3.2. Modeling approaches

The selected articles rely on mathematical modeling for analysis, simulation, and prediction of the association between human mobility and COVID-19. According to Siettos et al.’s categories of mathematic modeling of infectious disease dynamics, we categorized the mathematical models used in the selected articles into three categories: statistical method, mathematical/mechanistic state-space model, and simplified arithmetic model (Siettos and Russo 2013). The category of these analytical models was presented in Figure 2.

**Figure 2.**
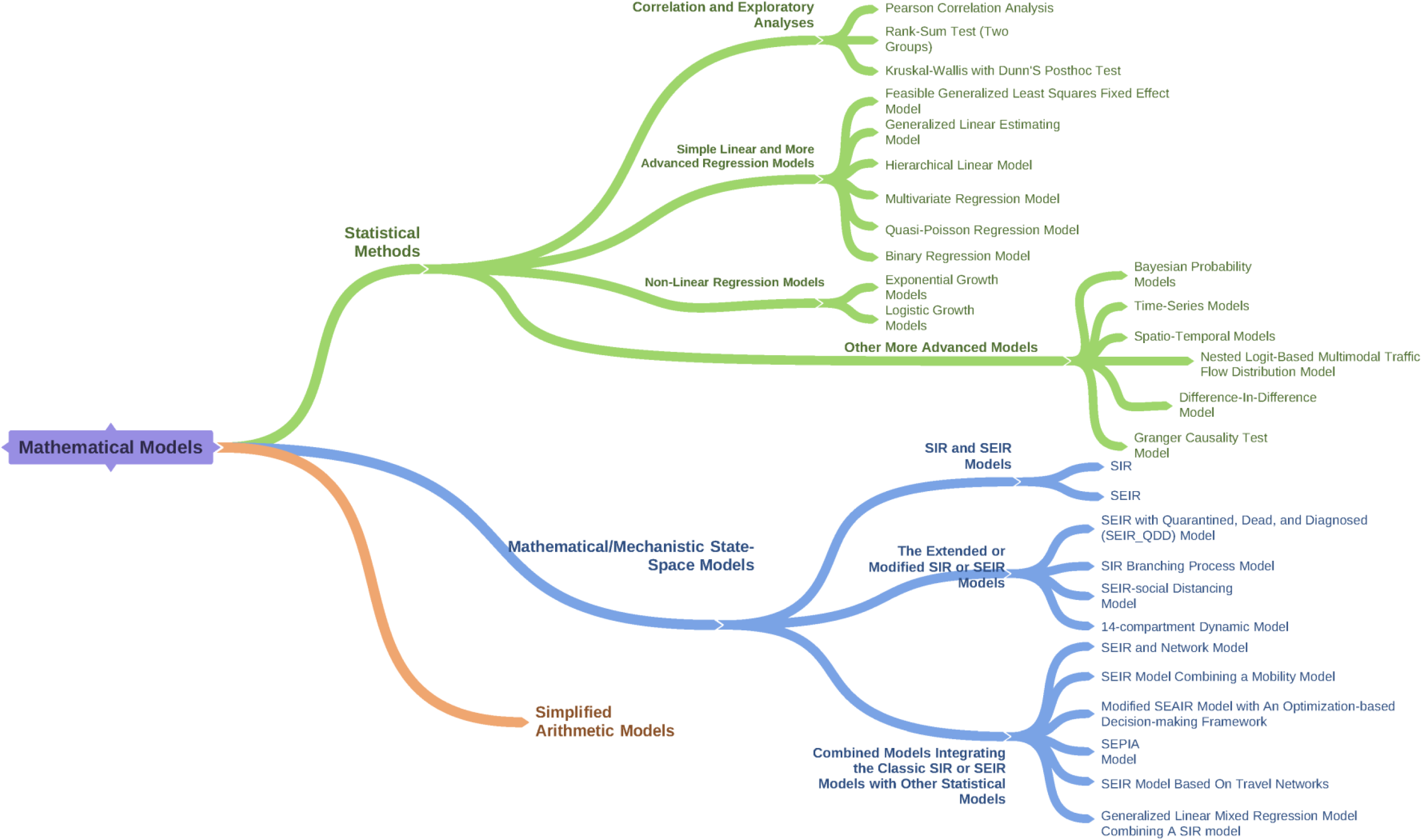
Summary of mathematical models applied in the selected articles

A total of 26 selected articles apply statistical methods, including correlation and exploratory analyses, as well as simple linear and advanced regression models. These models are mainly used in evaluating the effectiveness of social restriction policies and modeling the relationship between human mobility and COVID-transmission. The correlation and exploratory analyses conducted in these articles consist of Pearson correlation analysis (Chen, Chen et al. 2020, Zhang, Chen et al. 2020), rank-sum test (two groups) (McGrail, Dai et al. 2020), and Kruskal-Wallis test with Dunn’s post hoc analysis (McGrail, Dai et al. 2020). Other studies employ simple linear regressions models, including feasible generalized least squares fixed effect model (Yang, Chen et al. 2020, Zhang, Litvinova et al. 2020), generalized linear estimating model (Kraemer, Yang et al. 2020), hierarchical linear model (Alfano and Ercolano 2020), multivariate regression model (Sirkeci and Yüceşahin 2020), quasi-Poisson regression model (Tobías 2020), and binary regression model (Yuan, Xiao et al. 2020). However, such methods have limitations; for example, the variables in most of these models should be linear and independent from each other. If the relationship between the variables is non-linear, it may produce misleading inferences. To address the issue of non-linearity and endogeneity issues, some researchers applied non-linear regression models, including exponential growth models (Courtemanche, Garuccio et al. 2020, Djurović 2020) and logistic growth models (Aviv-Sharon and Aharoni 2020, Zou, Pan et al. 2020). Another stream of studies uses more advanced models to predict the future trend of COVID-19, including Bayesian probability models (Bherwani, Anjum et al. 2020, Kaur, Bherwani et al. 2020), time-series models (Jia, Lu et al. 2020, Jiang and Luo 2020, Moosa 2020, Salje, Tran Kiem et al. 2020), spatial-temporal models (Dickson, Espa et al. 2020, Jia, Lu et al. 2020, Tian, Liu et al. 2020), nested logit-based multimodal traffic flow distribution model (Zheng 2020), difference-in-difference model (Banerjee and Nayak 2020), and granger causality test model (Zhao, Wang et al. 2020).

Mathematical/mechanistic state-space models (dynamic system models) were used in 21 selected articles. These models are commonly used in epidemiological studies to stimulate or predict the future trend of COVID-19 transmission with the inclusion of human mobility data as model parameters or other types of data (e.g., road network, anti-body) as variables to optimize the modeling performance. Among those, Susceptible-Infectious-Recovered (SIR) and Susceptible-Exposed-Infectious-Recovered (SEIR) models (Balmford, Annan et al. 2020, DeFries, Agarwala et al. 2020, Neufeld, Khataee et al. 2020, Rainisch, Undurraga et al. 2020, Roda, Varughese et al. 2020) are the most widely used models. Both SIR and SEIR models are used to predict the number of confirmed, susceptible, recovered cases, and deaths with the involvement of human mobility measures as variables or modeling parameters. Other studies employ the extended or modified SIR or SEIR models to improve the model performance (Ding and Gao 2020, Ngonghala, Iboi et al. 2020, Sun, Wang et al. 2020, Yang, Qi et al. 2020, Zhang, Dong et al. 2020), including SEIR with Quarantined, Dead, and Diagnosed (SEIR-QDD) model (Wang, Xu et al. 2020), SIR branching process model (O’Sullivan, Gahegan et al. 2020), SEIR-social distancing model (Gupta, Jain et al. 2020), and a 14-compartment dynamic model (Westerhoff and Kolodkin 2020). The remaining articles utilize combined models integrating the classic SIR or SEIR models with other statistical models, including SEIR and network model (Peirlinck, Linka et al. 2020), SEIR model combining mobility model (Linka, Peirlinck et al. 2020), modified SEAIR model with optimization-based decision-making framework (Tsay, Lejarza et al. 2020), SEPIA model (Gatto, Bertuzzo et al. 2020), SEIR model based on travel networks (Lai, Ruktanonchai et al. 2020), and generalized linear mixed regression model combining SIR model (Zhang, Litvinova et al. 2020).

Among the selected articles, one study uses a simplified arithmetic model (Killeen and Kiware 2020) with simple calculations (e.g., addition, subtraction, multiplication, division, rounding off, a few conditional statements, and two unavoidable power terms) to ease the interpretability of the model. This model enables non-specialist readers to understand the process of modeling and in-depth inspect numerical predictions.

### 3.3. Study purposes and key findings

The selected articles aim to 1) examine the effectiveness of policy-induced mobility control on COVID-19 (hereinafter referred to “policy implementation and evaluation”); 2) predict the COVID-19 dynamic through modeling or simulating human mobility or its related measures (hereinafter referred to “simulation and prediction”); 3) compare studies presenting the association between human mobility and COVID-19 across countries or regions (hereinafter referred to as “cross-country/region comparison”). The key findings of each purpose were presented below. Some articles may fall into more than one category if they contribute to each category equivalently.

#### 3.3.1. Policy implementation and evaluation

The primary purpose of the selected articles focuses on estimating the influence of policy-induced human mobility on the transmission of COVID-19, particularly through assessing mobility change caused by social distancing, lockdown orders, and travel restrictions.

Some common findings from these policy-oriented papers show that policy interventions including lockdown, travel restrictions, social distancing, and border control have effectively reduced the transmission of COVID-19 (Chen, Chen et al. 2020, Djurović 2020, Jiang and Luo 2020, Tian, Liu et al. 2020, Yang, Chen et al. 2020, Yang, Qi et al. 2020). Articles focusing on the experience in Wuhan in China, where the first COVID-19 case was reported, found that the lockdown implemented in Wuhan substantially mitigated the spread of COVID-19 and delayed the growth of the COVID-19 epidemic in other cities in Hubei and other provinces in China (Chen, Chen et al. 2020, Jiang and Luo 2020, Kraemer, Yang et al. 2020, Tian, Liu et al. 2020, Yang, Chen et al. 2020, Zhang, Chen et al. 2020). Moreover, some studies give an appraisal on China’s response to COVID-19 that the lockdown strategy adopted by the Chinese government and the launching of the first-level response to COVID-19 have had prompt, timely, and positive effects on controlling the pandemic (Chen, Chen et al. 2020, Jiang and Luo 2020, Sun, Wang et al. 2020, Yang, Chen et al. 2020). Similarly, in Europe, the effectiveness of lockdown policies on control the spread of COVID-19 has also been witnessed in Italy, Spain, and France (Dickson, Espa et al. 2020, Salje, Tran Kiem et al. 2020, Tobías 2020). Mobility restriction measures implemented in the U.S. also effectively decreased the spread of COVID-19. At the same time, researchers were concerned about the potential danger of an exponential spread of the virus in the absence of interventions as the ongoing debates of the necessity and effectiveness of social distancing in the U.S. government and the society(Banerjee and Nayak 2020, Courtemanche, Garuccio et al. 2020).

The relationship between human mobility and the virus spread is temporal and spatial heterogeneity, along with observing a time-lag effect of mobility on the virus spread. Policy interventions, despite being globally effective in reducing both the spread of infection and its self-sustaining dynamics, have had heterogeneous impacts locally (Dickson, Espa et al. 2020, O’Sullivan, Gahegan et al. 2020, Zhang, Chen et al. 2020). For example, large metropolitan areas encounter more disruptions and larger challenges to control infection because they cannot easily be broken down into separately managed regions (O’Sullivan, Gahegan et al. 2020). Labor-intensive cities in China need to take stronger measures to prevent a potential rebound in COVID-19 cases after releasing the restriction policies (Zhang, Chen et al. 2020). Lockdown on public transport (e.g., auto, railway, coach, and flight) in China has the most prominent impact on virus control compared to lockdown on other public spaces (Zheng 2020). Researchers found in India that a prudent post-lockdown strategy might focus on easing physical distancing restrictions within high-risk places while maintaining restrictions between high-risk places (DeFries, Agarwala et al. 2020) Moreover, policy measures need to be adjusted across the different phases of the pandemic. In the initial stage of the outbreak, human mobility from Wuhan to other places in China was highly relevant to the growth rate of the COVID-19 cases in other cities and provinces. Still, this association became negative after the implantation of Wuhan lockdown and other travel restrictions nationally (Kraemer, Yang et al. 2020, Zhang, Chen et al. 2020). Additionally, the reduction of infection caused by mobility control is observed to be relatively weaker in places where the outbreak occurred later (Zhang, Chen et al. 2020).

Furthermore, mobility control is observed to have a time-lag effect on the virus transmission and such effect varies across the geographic contexts and the timeline of the pandemic. In this U.S., researchers found that social distancing reduced the daily growth rate of confirmed COVID-19 cases by 5.4 percentage after one to five days but 9.1 percentage points after sixteen to twenty days (Courtemanche, Garuccio et al. 2020). Studies across various countries reported that the efficacy of lockdown continues to hold over two weeks or even up to 20 days after a lockdown was implemented (Alfano and Ercolano 2020, McGrail, Dai et al. 2020).

Scholars find that the timing, effectiveness, and stringency of policy implementation are crucial for the success of COVID-19 control efforts in different countries (Gupta, Jain et al. 2020, Ngonghala, Iboi et al. 2020, Sun, Wang et al. 2020)(). The early implementation of social and mobility restrictions is especially effective in lowering the peak value of new infections and reducing the infection scale (Bherwani, Anjum et al. 2020, Kaur, Bherwani et al. 2020, Sun, Wang et al. 2020, Yuan, Xiao et al. 2020, Zhang, Chen et al. 2020). Ngonghala et al. asserted that ensuring the high adherence/coverage of policy intervention and enhancing the effectiveness of such intervention is particularly important in control infection in the local community(Ngonghala, Iboi et al. 2020). However, policymakers are more concerned about the public pressure towards lockdown mitigation as well as the downside of restrictive lockdown, for example, the tradeoff of social and economic upheaval (Westerhoff and Kolodkin 2020). For example, Tsay et al. support the “on-off” policies alternating between strict social restriction and relaxing such restrictions can be effective at flattening the infection curve while likely minimizing social and economic cost, especially for the places where persistent small outbreaks oscillate between high-risk regions for many months (Tsay, Lejarza et al. 2020).

#### 3.3.2. Simulation and prediction

Another stream of the selected articles focuses on proposing and developing mathematical models to simulate and predict dynamics of COVID-19 with various assumed plans or policies in limiting human mobility or altering time variances of certain plan and policies to predict disease changes (e.g.,(Gupta, Jain et al. 2020, Jia, Lu et al. 2020, Linka, Peirlinck et al. 2020)).

Specifically, these quantitative modeling work aim to quantify the COVID-19 pandemic by various mathematical/mechanistic state-space models as summarized in the modeling section (section 3.2), including epidemic models (Aviv-Sharon and Aharoni 2020, Ding and Gao 2020, Djurović 2020, Gatto, Bertuzzo et al. 2020, Lai, Ruktanonchai et al. 2020, Peirlinck, Linka et al. 2020, Roda, Varughese et al. 2020, Sirkeci and Yüceşahin 2020, Wang, Xu et al. 2020, Zhao, Wang et al. 2020), spatial-temporal models (Bherwani, Anjum et al. 2020, Jia, Lu et al. 2020, O’Sullivan, Gahegan et al. 2020), biological models (Westerhoff and Kolodkin 2020), and other advanced mathematical models (Killeen and Kiware 2020, Tsay, Lejarza et al. 2020, Yang, Qi et al. 2020). The common characteristic of these modeling approaches involves the measures of human mobility and social restriction policies as parameters into the modeling configuration.

The modeling work conducted in our selected papers provides similar findings regarding policy implementation and mobility control as described above. Also, these modeling work bring in technical benefits as follows to help estimate outbreak dynamics and provide decision guidelines for successful outbreak control (Peirlinck, Linka et al. 2020). First, policy interventions parameterized in the modeling process are adjustable, allowing the evaluation of local policy scenarios and relaxing political measures (Rainisch, Undurraga et al. 2020). For example, Gatto et al. measured averted hospitalizations by running scenarios obtained by selectively relaxing the imposed restrictions to support the planning of emergency measures (Gatto, Bertuzzo et al. 2020). Second, developing multi-disciplinary models are able to explore temporal changes in spreading patterns and outbreak dynamics (Linka, Peirlinck et al. 2020) and estimate the potentials of vaccination (Peirlinck, Linka et al. 2020). As Salje et al. asserted, population immunity appears insufficient to avoid a second wave if all social restrictions are released at the end of the lockdown (Salje, Tran Kiem et al. 2020). Third, some models can predict the outbreak spreading and the pandemic cessation dates (Aviv-Sharon and Aharoni 2020, Jia, Lu et al. 2020). The comparison of modeling results across different locales and various scenarios serves as a direct tool to detect community transmission burden (Jia, Lu et al. 2020).

#### 3.3.3. Cross-country/region comparison

Another important purpose of the selected articles is to compare policy implementation responding to the pandemic, economic and financial consequences of lockdown orders, and price of life comparisons across countries, regions, and cities (Alfano and Ercolano 2020, Aviv-Sharon and Aharoni 2020, Balmford, Annan et al. 2020, Gul, Tuncay et al. 2020, Linka, Peirlinck et al. 2020, McGrail, Dai et al. 2020, Moosa 2020, Yang, Qi et al. 2020). Such studies provide empirical evidence on the influence of human mobility on the COVID-19 in 8 countries (Balmford, Annan et al. 2020), 10 countries (Moosa 2020), across European countries (Linka, Peirlinck et al. 2020), across Asian countries (Aviv-Sharon and Aharoni 2020), and between Wuhan in China and London in the UK (Yang, Qi et al. 2020).

Several comparison studies reveal findings specific to different geographic contexts that have not been covered in the previous summary. In general, policy interventions may well explain the majority of cross-country variation in virus control in the initial stage of the pandemic (Balmford, Annan et al. 2020). However, these are less definitive conclusions if extended to a full spectrum of the pandemic. Mobility restriction policies implemented during the pandemic differ widely around the world. Policies that work well in one country may not be effective in other places. For instance, Kaur et al. indicated that countries that acted late in bringing in the policy intervention suffered from a higher infection rate than countries that reacted faster (Kaur, Bherwani et al. 2020). It is partially in line with the findings from a 10-country comparison that countries that have not imposed lockdown or have imposed lockdown either late or without stringency have performed poorly in infection control, except for Korea (Moosa 2020). The outbreak in Korea has been controlled rather well without a full lockdown, as Korea conducted a combination of interventions including border control, testing, tracing, the quality of the healthcare system, preparedness for epidemics and pandemics, and population density (Moosa 2020). Thus, when it comes to implementing different policy approaches to the pandemic, careful consideration of cross-country differences is required in terms of countries’ nature as well as their demographic and socioeconomic variations. Yang et al. observed that China has efficient government initiatives and effective collaborative governance for mobilizing corporate resources to provide essential goods; however, this mode may be not suitable to the UK where it is more possible to take a hybrid intervention of suppression and mitigation to balance the total infections and economic loss (Yang, Qi et al. 2020).

## 4. Discussion: future directions and limitations

Based on the preceding systematic review and responding to the remaining concerns mentioned in the selected articles, we call for concerted attention of the COVID-19 research community to the following directions: 1) to encourage multi-disciplinary collaboration with joint efforts from researchers with different backgrounds; 2) to adjust the implementation and stringency of mobility-control policies flexibly in correspond to the rapidly changing trend of COVID-19; 3) to improve the methods used in analyzing, simulating, and predicting COVID-19 to be more realistic, context-specific, and temporal-specific; and 4) to enrich mobility data sources as well as improve data accuracy and suability for applications.

### 4.1. Multidisciplinary collaboration

Future studies should involve the expertise of researchers and professionals across disciplines. First, since covert transmission may be the main mode of spread and subject to the risk of a second epidemic peak, and its role is severely underestimated (Ding and Gao 2020), epidemiologists and other health professionals can contribute to the measurement of covert cases, which can be used in future modeling. Second, policy interventions that are widely discussed to control mobility are lockdown, social distancing, and travel restrictions. However, these strategies can be extended to border controls at a regional or national level as well as testing and contact tracing policies at an individual level (Moosa 2020). Herein, policymakers and stakeholders should be involved to optimize the quantification of policies. Third, the effect of seasonality on the transmission dynamics of COVID-19 is underestimated in current scholarship (Alfano and Ercolano 2020, Gupta, Jain et al. 2020), although some scholars assume that the decrease of COVID-19 infection may be suspiciously attributable to unknown seasonal factors, for example, temperature and absolute humidity (Lai, Ruktanonchai et al. 2020, Rainisch, Undurraga et al. 2020, Zhang, Chen et al. 2020). Referring to the fact that the transmission of similar respiratory illnesses (e.g., influenza, syncytial virus) peaks in winter (Lipsitch and Viboud 2009, Shaman and Kohn 2009), the evaluation of the seasonality effect is needed.

### 4.2 Policy adjustment

With an increasing number of countries experiencing the second wave of the pandemic, further work is needed to determine how to optimally balance the trade-off between economic loss and health outcomes of COVID-19 (Kraemer, Yang et al. 2020). Policy interventions have been gradually upgraded along the timeline of the pandemic, which greatly promotes the arrival of the turning point of the epidemic (Jiang and Luo 2020). As such, more evaluations about the effectiveness of intermediate measures become important to control the social and economic cost, such as lifting a shelter-in-place order but requiring masks in public or opening restaurants at reduced capacity (Ngonghala, Iboi et al. 2020). There is also a need to include other factors in prevention and control measures, such as case detection, contact tracing, quarantining, quality of healthcare systems, and population and housing density (Gupta, Jain et al. 2020, Moosa 2020). The resurge of infection has also been observed to be associated with the release of national border controls (Moosa 2020); therefore, widespread decisive national action and international co-operation are required to conditionally reopen trade and travel between countries. Great caution is needed as gradual, exploratory steps toward reopening (Courtemanche, Garuccio et al. 2020) as even a slight relaxation of lockdown or importation controls may cause containment failure (Killeen and Kiware 2020). A combination of multiple interventions may achieve the strongest and most rapid effect on containing the spread of the virus (Lai, Ruktanonchai et al. 2020). In addition, health education about the risk and severity of COVID-19 infection is needed to increase public awareness (Ding and Gao 2020).

### 4.3. Methodological improvement in spatial and temporal dimensions

Future research is needed to improve the accuracy of Epidemiological models (e.g., dominantly SEIR models) by involving additional COVID-19 related information, such as the number of asymptomatic cases (Lai, Ruktanonchai et al. 2020, Peirlinck, Linka et al. 2020) and the incubation period of the novel coronavirus (Jiang and Luo 2020). Model parameters can involve the measures of the effectiveness of policy implementation (Gupta, Jain et al. 2020) and pharmaceutical factors (e.g., improved medical treatments, vaccine development, viral mutation, increased likelihood of testing for subjects with more severe symptoms, the probability of changing antigenicity and virulence (Tsay, Lejarza et al. 2020), as well as the quantification of other non-pharmaceutical factors that are likely to contribute to control virus work, especially the isolation of suspected and confirmed patients and their contact (Tian, Liu et al. 2020, Zhang, Dong et al. 2020). For more hybrid modeling work across disciplines, individual-level models need to include many patient-specific factors, including demographic and socioeconomic status (e.g., age-sex structure, ethnicity, and income) (Zhang, Chen et al. 2020, Zhang, Dong et al. 2020). Aggregated-level models can extend to consider area-specific factors to distinguish heterogeneity within the regions (Alfano and Ercolano 2020, Dickson, Espa et al. 2020), including geographical and spatial characteristics (e.g., location, population, and housing density in a suburb) (Moosa 2020, Zhang, Dong et al. 2020) given the built environment in neighborhoods where confirmed or suspected cases reside would affect the likelihood of infection (Gupta, Jain et al. 2020, O’Sullivan, Gahegan et al. 2020). Collectively, further research can be carried out in unifying temporary and spatial dimensions by distinguishing the different stages of pandemic and involving time-dependent parameters for a holistic understanding of the infection risk at hand (Bherwani, Anjum et al. 2020, Peirlinck, Linka et al. 2020, Sun, Wang et al. 2020).

### 4.4 Enrichment of spatial-temporal data from multiple sources

Migration data used in current publications largely came from public sources, including data released by large companies (e.g., migration data from Baidu, Google, Apple, Facebook, and Twitter), global mobility network, and mobile phone companies (Zhang, Dong et al. 2020). There are limitations of these data sources. Some data are country-specific; for example, the Baidu migration data is only available in China (Yuan, Xiao et al. 2020). Second, mobility data retrieved from mobile phones or mobile app users designed by large companies encounter data biases in population coverage, which may exclude some specific subgroups particularly children and aged populations who may not use mobile phones (Banerjee and Nayak 2020, Lai, Ruktanonchai et al. 2020). The index-based mobility data (e.g., provided by Google, Baidu, and Apple) does not include population inflow to and/or outflow from a given place. Alternatively, user-based social media big data (e.g., geotagged Twitter data) is able to indicate the inter-regional movement to improve the accuracy of models (Gupta, Jain et al. 2020, O’Sullivan, Gahegan et al. 2020, Tsay, Lejarza et al. 2020), although such big data is less used in current studies. With the technological advancements and the emergence of further refined data, it will be interesting in future studies to involve additional data, to use a combination of multi-sourced data, and to compare the reliability and quality of data (Banerjee and Nayak 2020, McGrail, Dai et al. 2020). Moreover, data sharing and information disclosure are encouraged for future studies. Some scholars and institutes have put great efforts into collecting, collating, and sharing data via crowdsourcing and cloud platforms to facilitate cross-disciplinary collaborations. For example, Harvard Dataverse provides an open online data management and sharing platform for COVID-19 studies with daily COVID-19 confirmed cases, global news, social media data, population mobility, climate, health facilities, socioeconomic data, events chronicle, and scholarly articles (Hu et al., 2020).

### 4.5 Limitation

The study has limitations that should be noted. First, we did not include non-peer-reviewed articles (e.g., working papers and preprints) in this review. Traditional peer review usually takes months from submission to publication, while timely reporting of research findings is a priority during the pandemic, which dramatically increased the use of preprint service (Jung, Sun et al. 2020). Though preprints provide direct and rapid access to information, criteria used to justify preprints are not available. Thus, we only searched for published and early access articles, which inevitably exclude the findings from some popular non-peer-reviewed articles. Second, this review includes a small number of eligible articles focusing on Africa and South America which could be due to the late appearance of the first case in some regions as well as the limited funding and resource to conduct COVID-19 related research. Third, our search obtained limited articles covering the second and third waves of the COVID-19 pandemic, which has been observed in several countries after lifting mobility restriction policies. The findings summarized in this review may not well explain the resurged cases or the cases via converted transmission over a long time. Thus, we encourage future researchers to extend our systematic review to cover a longer period and to include the most updated results from both published and preprint articles from various regions.

## 5. Conclusion

Understanding the pattern of human mobility is essential to prevent and predict the spread of infectious diseases. As COVID-19 continues to spread and resurge across countries, we summarized data and analytical models used in publications related to human mobility and COVID-19 transmission. The authors applied various models including statistical models, mathematical/mechanistic state-space models, and simplified arithmetic models to examine the relationship between human mobility and COVID-19 transmission, using multi-sourced spatial-temporal mobility data. The findings on policy implications summarized herein provide important guidance in making, implementing, and adjusting current and post-pandemic measures. What we have seen in existing studies is the relationship between human mobility and the virus spread is temporal and spatial heterogeneity, along with the observation of a time-lag effect of mobility on the spread of the virus. Additionally, this relationship is stronger in the initial stage of the pandemic but less conclusive if extends to a full spectrum of the pandemic or different geographic contexts. What we have not seen from the current publications motivates us to propose future research directions. Specifically, we suggested that governments promote prompt and sustainable measures to control the spread of COVID-19. We also encourage multi-disciplinary collaborators to conduct rapid and accurate risk assessments of the pandemic by incorporating rich data sources and improving spatial-temporal modeling to prevent and predict future outbreaks.

## Data Availability

N/A

## Acknowledgment

This study is supported by the National Science Foundation [1841403; 2027540].

